# Larger perivascular space volume fraction is associated with worse post-stroke sensorimotor outcomes: An ENIGMA analysis

**DOI:** 10.1101/2024.12.20.24319296

**Authors:** Stuti Chakraborty, Jeiran Choupan, Octavio Marin-Pardo, Mahir H. Khan, Giuseppe Barisano, Bethany P. Tavenner, Miranda R. Donnelly, Aisha Abdullah, Justin W. Andrushko, Nerisa Banaj, Michael R. Borich, Lara A. Boyd, Cathrin M. Buetefisch, Adriana B. Conforto, Steven C. Cramer, Martin Domin, A. Adrienne Dula, Jennifer K. Ferris, Brenton Hordacre, Steven A. Kautz, Neda Jahanshad, Martin Lotze, Kyle Nishimura, Fabrizio Piras, Kate P. Revill, Nicolas Schweighofer, Surjo R. Soekadar, Shraddha Srivastava, Sophia I. Thomopoulos, Daniela Vecchio, Lars T. Westlye, Carolee J. Winstein, George F. Wittenberg, Kristin A. Wong, Paul M. Thompson, Sook-Lei Liew

**Affiliations:** Chan Division of Occupational Science and Occupational Therapy, University of Southern California, Los Angeles, CA, USA; Department of Neurology, University of Southern California, Los Angeles, CA, USA; Neuroscience Graduate Program, University of Southern California, Los Angeles, CA, USA; Department of Neurosurgery, Stanford University, Stanford, CA, USA; Department of Psychology, University of California Riverside, Riverside, CA, USA; Department of Occupational Therapy and Occupational Science; Department of Sport, Exercise and Rehabilitation, University of Northumbria, Newcastle upon Tyne, UK; Laboratory of Neuropsychiatry, IRCCS Santa Lucia Foundation, Rome, Italy; Division of Physical Therapy, Department of Rehabilitation Medicine, Emory University School of Medicine, Atlanta, GA; Department of Physical Therapy, University of British Columbia, Vancouver, BC, Canada; Department of Neurology, Emory University School of Medicine, Atlanta, GA, USA; Hospital das Clínicas, São Paulo University, São Paulo, Brazil; Hospital Israelita Albert Einstein, São Paulo, Brazil; Department of Neurology, David Geffen School of Medicine, University of California, Los Angeles, CA, USA; California Rehabilitation Institute; Los Angeles, CA, USA; Functional Imaging Unit, Department for Radiology and Neuroradiology, University Medicine Greifswald, Greifswald, Germany; Department of Neurology, Dell Medical School, The University of Texas at Austin, Austin, TX, USA; Gerontology Research Center, Simon Fraser University, Vancouver, British Columbia, Canada; Innovation, Implementation and Clinical Translation (IIMPACT) in Health, Allied Health and Human Performance, University of South Australia, Adelaide, Australia; Ralph H Johnson VA Health Care System, Charleston, SC, USA; Department of Health Sciences and Research, Medical University of South Carolina, Charleston, SC, USA; Imaging Genetics Center, Mark and Mary Stevens Neuroimaging and Informatics Institute, Keck School of Medicine, University of Southern California, Los Angeles, CA; Clinical Neuroscience and Neurorehabilitation, IRCCS Santa Lucia Foundation, Rome, Italy; Facility for Education and Research in Neuroscience, Emory University, Atlanta, GA, USA; Division of Biokinesiology and Physical Therapy, University of Southern California, Los Angeles, CA, USA; Clinical Neurotechnology Laboratory, Department of Psychiatry and Neurosciences (CCM), Charité - Universitätsmedizin Berlin, Berlin, Germany; Courage Kenny Rehabilitation Institute, Allina Health, Minneapolis, MN; Department of Psychology, University of Oslo, Norway, Oslo, Norway; Center for Precision Psychiatry, Division of Mental Health and Addiction, Oslo University Hospital, Oslo, Norway; Department of Neurology, Keck School of Medicine, University of Southern California, Los Angeles, CA, USA; Department of Neurology, University of Pittsburgh, Pittsburgh, PA, USA; Department of Veterans Affairs Geriatrics Research Educational & Clinical Center, Veterans Affairs Pittsburgh Healthcare System (VAPHS), Pittsburgh, PA, USA; Department of Physical Medicine & Rehabilitation, Dell Medical School, The University of Texas at Austin, Austin, TX, USA; Department of Biomedical Engineering, Viterbi School of Engineering, University of Southern California, Los Angeles, CA, USA

**Author notes:** **Correspondence to**: Sook-Lei Liew, PhD, OTR/L, FAOTA Chan Division of Occupational Science and Occupational Therapy, Division of Biokinesiology and Physical Therapy, Viterbi Department of Biomedical Engineering, Keck Department of Neurology, Stevens Neuroimaging and Informatics Institute, University of Southern California, 1540 Alcazar Street, Los Angeles, CA 90089, United States.

**Keywords:** stroke, global brain health, perivascular spaces, sensorimotor outcomes, cerebral small vessel disease, white matter hyperintensities, brain age

## Abstract

**BACKGROUND:** Perivascular Spaces (PVS) are a marker of cerebral small vessel disease (CSVD) that are visible on brain imaging. Larger PVS has been associated with poor quality of life and cognitive impairment post-stroke. However, the association between PVS and post-stroke sensorimotor outcomes has not been investigated.

**METHODS:** 602 individuals with a history of stroke across 24 research cohorts from the ENIGMA Stroke Recovery Working Group were included. PVS volume fractions were obtained using a validated, automated segmentation pipeline from the basal ganglia (BG) and white matter centrum semiovale (CSO), separately. Robust mixed effects regressions were used to a) examine the cross-sectional association between PVS volume fraction and post-stroke sensorimotor outcomes and b) to examine whether PVS volume fraction was associated with other measures of CSVD and overall brain health (e.g., white matter hyperintensities [WMHs], brain age [measured by predicted age difference, brain-PAD]).

**RESULTS:** Larger PVS volume fraction in the CSO, but not BG, was associated with worse post-stroke sensorimotor outcomes (b = -0.06, p = 0.047). Higher burden of deep WMH (b = 0.25, p <0.001), periventricular WMH (b = 0.16, p <0.001) and higher brain-PAD (b = 0.09, p <0.001) were associated with larger PVS volume fraction in the CSO.

**CONCLUSIONS:** Our data show that PVS volume fraction in the CSO is cross-sectionally associated with sensorimotor outcomes after stroke, above and beyond standard lesion metrics. PVS may provide insight into how the overall vascular health of the brain impacts inter-individual differences in post-stroke sensorimotor outcomes.

## INTRODUCTION

Measures that quantify the direct impact of the lesion after stroke, such as corticospinal tract lesion load (CST-LL), have been consistently related to post-stroke sensorimotor outcomes.^1,2^ However, there is still unexplained inter-individual variability in sensorimotor outcomes after stroke that is not fully captured by considering lesion measures such as CST-LL alone.^3,4^ Recent work suggests that measures of overall brain health may also explain inter-individual variability in post-stroke sensorimotor outcomes.^5^ These global brain health (GBH) measures focus on capturing the vascular, cellular, and structural integrity of the entire brain, using noninvasive methods such as Magnetic Resonance Imaging (MRI).^6,7,8^ Several measures of GBH include vascular markers of underlying cerebral small vessel disease (CSVD), like white matter hyperintensities (WMHs)^10^, and markers of structural brain integrity (e.g., brain age).^6^ To date, greater WMH burden has been associated with poorer functional outcomes after stroke,^11^ increased risk of dementia and cognitive impairment.^12^ Recently, larger WMH volume was also associated with worse post-stroke motor impairment and was shown to modify the relationship between motor impairment and CST-LL.^13^ Similarly, brain-PAD was associated with worse post-stroke sensorimotor outcomes and was shown to mediate the relationship between CST-LL and sensorimotor outcomes.^6^

Perivascular spaces (PVS), another measure of whole-brain health, may also impact sensorimotor outcomes. PVS are fluid-filled compartments that form a network around penetrating cerebral small vessels.^14,15^ PVS are an integral part of the glymphatic fluid clearance pathway of the brain and are essential for the clearance of metabolic waste products and interstitial solutes.^16,17^ Clearance of metabolic waste is especially important after stroke when necrotic tissue and other cellular debris must be removed to prevent further damage to brain tissue. While a few small PVS may be visible in the brain MRIs of young, healthy individuals, PVS become more visible when they are enlarged (e.g., 2 mm or greater).^18,19,20^ PVS enlargement may be indicative of poor metabolic waste clearance and blood-brain barrier dysfunction.^14,21^ Enlarged PVS commonly appear around the basal ganglia (BG) and white matter centrum semiovale (CSO) (Figure I) and are an imaging-based marker of underlying age-related CSVD.^1^

**Figure I:**
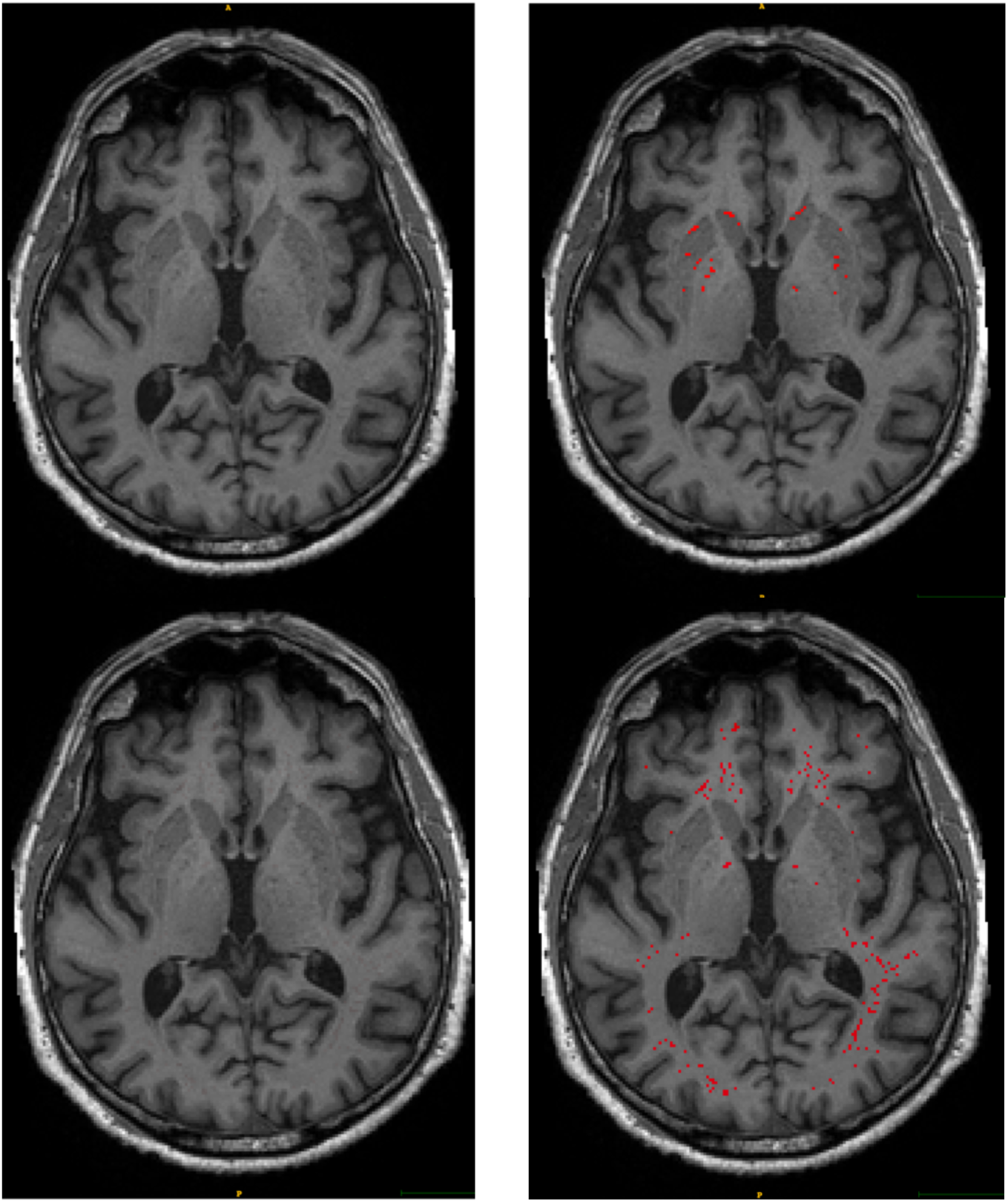
PVS in the basal ganglia (BG) and centrum semiovale (CSO) in stroke brain MRIs. Figure I: Axial 3D-T1 weighted slice of brain MRIs of individuals with a history of stroke. Left panel: brain slices showing underlying anatomy; Right panel: brain slices showing PVS segmented in red in the BG (top) and CSO (bottom).

CSVD is, in turn, associated with an increased risk of stroke or vascular dementia.^22^ Several risk factors have been associated with PVS enlargement, with old age being the most common demographic risk factor. Studies have also found biological male sex to be associated with PVS enlargement, more strongly in the CSO than in the BG.^20,23,24,2^ In relation to post-stroke outcomes, studies have reported an association between enlarged PVS and cognitive impairment at one year after stroke,^25^ lower quality of life^26^ and post-stroke depression.^27^ However, the relationship between PVS and sensorimotor outcomes after stroke has not been investigated.

In this study, we used PVS volume fraction obtained from a validated, automated segmentation pipeline to test the cross-sectional association between PVS and post-stroke sensorimotor outcomes, after controlling for other covariates. We hypothesized that larger PVS volume fraction would be associated with worse post-stroke sensorimotor outcomes. Further, we explored whether other GBH measures of vascular and structural integrity (such as WMH and brain-PAD, respectively), along with key demographic variables such as age, sex and time since stroke, were associated with PVS volume fraction.

## METHODS

### Data Availability Statement

The primary and corresponding authors of this study had full access to the data and take responsibility for data integrity and analyses. Publicly sharing the full data is limited by data sharing restrictions imposed by some of the (i) ethical review boards of the participating research cohorts, and consent documents; (ii) national and trans-national data sharing laws; and (iii) institutional processes, which may require a signed data transfer agreement for limited and predefined data use. However, similar to other ENIGMA working groups, we support data sharing among members of the ENIGMA Stroke Recovery Working Group who submit an analysis plan for a secondary project for group review. Following approval of the analysis plan, access to the relevant data will be provided, contingent on data availability, local PI approval and compliance with all supervising regulatory boards.

### Study Design and Data

This study used cross-sectional, pooled, multi-site data compiled by the ENIGMA Stroke Recovery Working Group, an international consortium of researchers that collects retrospective neuroimaging and behavioral data related to stroke.^28^ Detailed descriptions of the methods and procedures used by the ENIGMA Stroke Recovery Working Group and the datasets it has produced are published elsewhere.^28,29^ Data for this analysis was frozen on March 1, 2024. For this study, we queried the database for subjects with the following data elements: 1) a 3D T1-weighted MRI, 2) a manually segmented stroke lesion mask,^4^ 3) PVS segmentations generated by a validated, automated segmentation pipeline,^5^ and 4) an upper extremity sensorimotor outcome score.^6,7^ In addition, the following demographic information was extracted for each subject: sex, age, research cohort, and time since stroke (at the time of MRI). Ethical approvals for the data used in this study were collected in accordance with the Declaration of Helsinki and in compliance with the local ethics boards at each institute. Written informed consent was obtained from all study participants.

### Sensorimotor Outcome Score

In line with our previous work, we harmonized different sensorimotor outcome measures reported by each research cohort to generate a single *primary sensorimotor outcome score*, calculated as the percentage of the maximum possible score achieved by each subject (where 0 indicates severe impairment and 100 indicates no impairment).^30,6^ Details of the specific sensorimotor outcome measures used to calculate the sensorimotor outcome scores are reported by research cohort in the *Supplementary Material* (Supplementary Table I).

### MRI Data Analysis

Freesurfer (v.5.3.0)^31,32^ recon-all was used to segment T1-weighted MRIs and extract the total intracranial volume (ICV). Manual segmentation of stroke lesions was performed by trained research assistants, following a previously published ENIGMA tracing protocol.^28,33^ All stroke lesions were preprocessed by applying a registration to a standard MNI-152 template. Intensity nonuniformity correction was applied to account for bias field inhomogeneities; brain extraction using FMRIB Software Library (FSL BET) (FSL v.6.0.5) was performed to produce a skull-stripped brain mask.^29^ Affine linear transformation (with 12 degrees of freedom) was then applied using FSL’s FLIRT (FMRIB’s Linear Image Transformation Tool)^8^ function, with the brain mask as the cost function weighting mask to ensure outside structures do not influence the registration. The outputs obtained from skull stripping and affine linear transformation were visually inspected to ensure good quality brain segmentation and registration to the MNI template. We obtained stroke lesion volumes and the percent of CST-LL overlap using the Pipeline for Analyzing Lesions after Stroke (PALS) toolbox.^34^ A publicly available corticospinal tract (CST) template called the sensorimotor area tract template (SMATT), with 2x2x2 mm voxel resolution, was used to calculate percent of CST-LL overlap.^3^

### PVS Segmentation

The PVS segmentation algorithm was trained on T1-and T2-weighted images from the Human Connectome Project (HCP) S900 Release which included 900 healthy participants aged between 22-37 years and validated against a gold standard manual PVS visual rating scale (details published elsewhere).^36,37,38,39^ PVS was segmented from the BG and CSO separately, using only a T1-weighted image. Specific features of interest such as PVS counts, volumetric and spatial PVS distributions were obtained for each individual subject.^39^ PVS visibility was improved by removing non-structured noise; PVS volumes significantly corresponded to manual PVS ratings and manual PVS counts (see *Supplementary Materials*). For this analysis, only the PVS volume extracted from the pipeline was used. PVS volume fraction was then calculated for each region (BG, CSO). PVS volume fraction was calculated as the total volume of the segmented PVS voxel clusters in a specific region, divided by the total volume for that region. The final PVS volume output was obtained as:

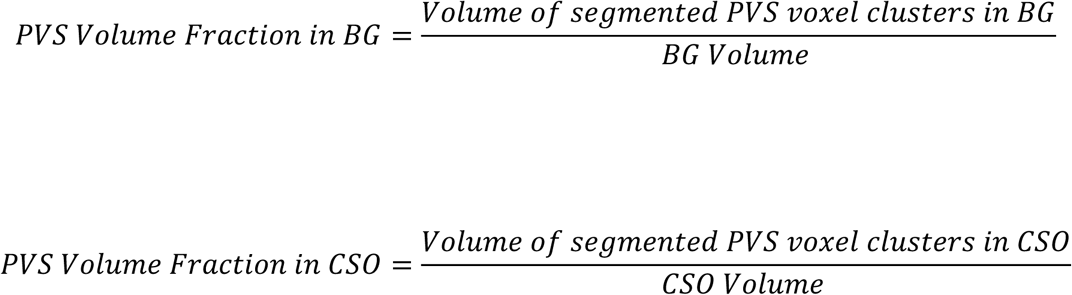

### Deep, Periventricular WMH Scores and Brain-PAD Calculation

We extracted other GBH measures such as deep and periventricular white matter hyperintensities (WMHs) and brain-PAD. Deep and periventricular WMHs were manually counted as lesions appearing on T1-weighted images.^40^ Visual estimation-based rating was done by an expert physician (G.B.) using Fazekas scores, where a score of 0 indicated no WMHs; 1 indicated between 1 – 4 WMHs; 2 indicated between 5 – 9 WMHs; and 3 indicated >9 WMH lesions.^41,42^

To calculate brain-PAD, we first obtained the predicted brain age using an openly available, previously published ridge regression model.^43^ We then calculated brain-PAD by subtracting the subject’s chronological age from the predicted brain age, as done previously in stroke.^6^ Figure II summarizes the data processing pipeline.

**Figure II:**
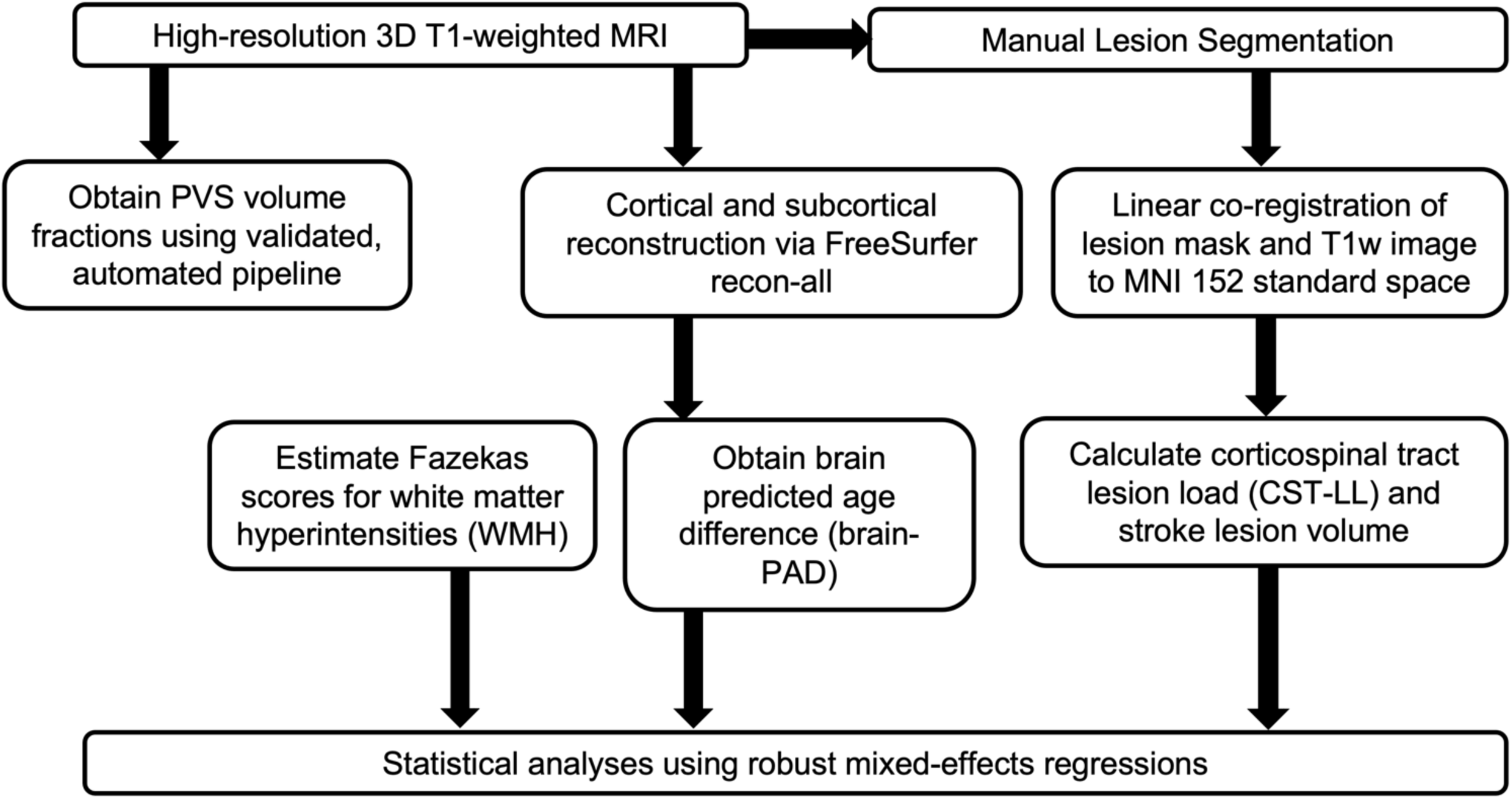
Summary of the Data Processing Pipeline. Details of the data processing pipeline

### Statistical Analyses

Statistical analyses were conducted in R (version 4.3.3, R core Team, 2021).^9^ We identified outliers in the data. To reduce the weight of these outliers without removing them, we used robust linear mixed-effects regression models.^10^ All variables were scaled. We examined the variance inflation factor (cut-off ≤ 2.5) for each fixed effect covariate included in our regression models to rule out the possibility of multicollinearity and reduce the redundancy of model predictors. For the primary analysis, we examined the cross-sectional association between PVS volume fraction (from BG and CSO as separate models) and post-stroke sensorimotor outcomes, with age, sex, time since stroke, CST-LL, stroke lesion volume and ICV as fixed effects covariates and research cohort as a random effect (*Supplementary Material, Model 1*). All statistical analyses were considered significant at p<0.05.

For our secondary analysis, we examined the association between PVS volume fraction in the CSO and BG, respectively, with GBH measures (deep and periventricular WMHs, brain-PAD), along with age, sex, time since stroke, CST-LL, ICV, and stroke lesion volume as fixed effects covariates, and research cohort as a random effect (*Supplementary Material*, *Models 2 and 3*).

## RESULTS

Cross-sectional data from 602 individuals with stroke (224 females and 378 males) across 24 research cohorts met the inclusion criteria to be included in this study. 53 subjects were in the acute phase (≤7 days since stroke), 78 subjects were in the early subacute phase (>7 and ≤90 days since stroke), 66 were in the late subacute (>90 and ≤180 days since stroke), and 405 were in the chronic (>180 days since stroke) phase. Summary statistics of the data are reported in Table I. Details of specific sensorimotor outcome measures used to calculate the sensorimotor outcome score, demographic and research cohort characteristics are outlined in the Supplementary Table II.

**Table I:** Summary Statistics of PVS Volume Fraction, Demographic, Stroke Lesion and Global Brain Health Covariates.

| *Covariate* | *Median* | *IQR (range)* |
| --- | --- | --- |
| PVS-BG volume fraction (mm<sup>3</sup>) | 0.028 | 0.022 (0.002 – 0.083) |
| PVS-CSO volume fraction (mm<sup>3</sup>) | 0.021 | 0.020 (0.001 – 0.068) |
| Age (years) | 61 | 16 (21-75) |
| Time Since Stroke (days) | 490 | 1174.75 (1- 7493) |
| Stroke Lesion Volume (mm<sup>3</sup>) | 5922.5 | 32037 (15 - 285724) |
| Deep WMH Fazekas Score | 1 | 2 (0 - 3) |
| Periventricular WMH Fazekas Score | 1 | 1 (0 - 3) |
| Brain-PAD (years) | 1.535 | 16.768 (-28.479 – +34.835) |

Summary statistics for each fixed effects covariate used in the mixed-effects regression models. The sample size (*n*), median, interquartile (*IQR*) and ranges are reported without scaling. Abbreviations: PVS volume fraction in BG = perivascular space volume in the basal ganglia; PVS volume fraction in CSO = perivascular space volume in the white matter centrum semiovale; WMH = white matter hyperintensities; Brain-PAD = brain predicted age difference.

### Larger PVS Volume Fraction in the CSO is Associated with Worse Post-Stroke Sensorimotor Outcomes

Larger PVS volume fraction, specifically in the white matter CSO, was significantly associated with worse post-stroke sensorimotor outcomes (b = -0.06, 95% CI = -0.12 – - 0.00, p = 0.047). However, PVS volume fraction in the BG was not significantly associated with post-stroke sensorimotor outcomes (b = 0.07, 95% CI = -0.00 – 0.14, p = 0.094) (Table II).

**Table II:**
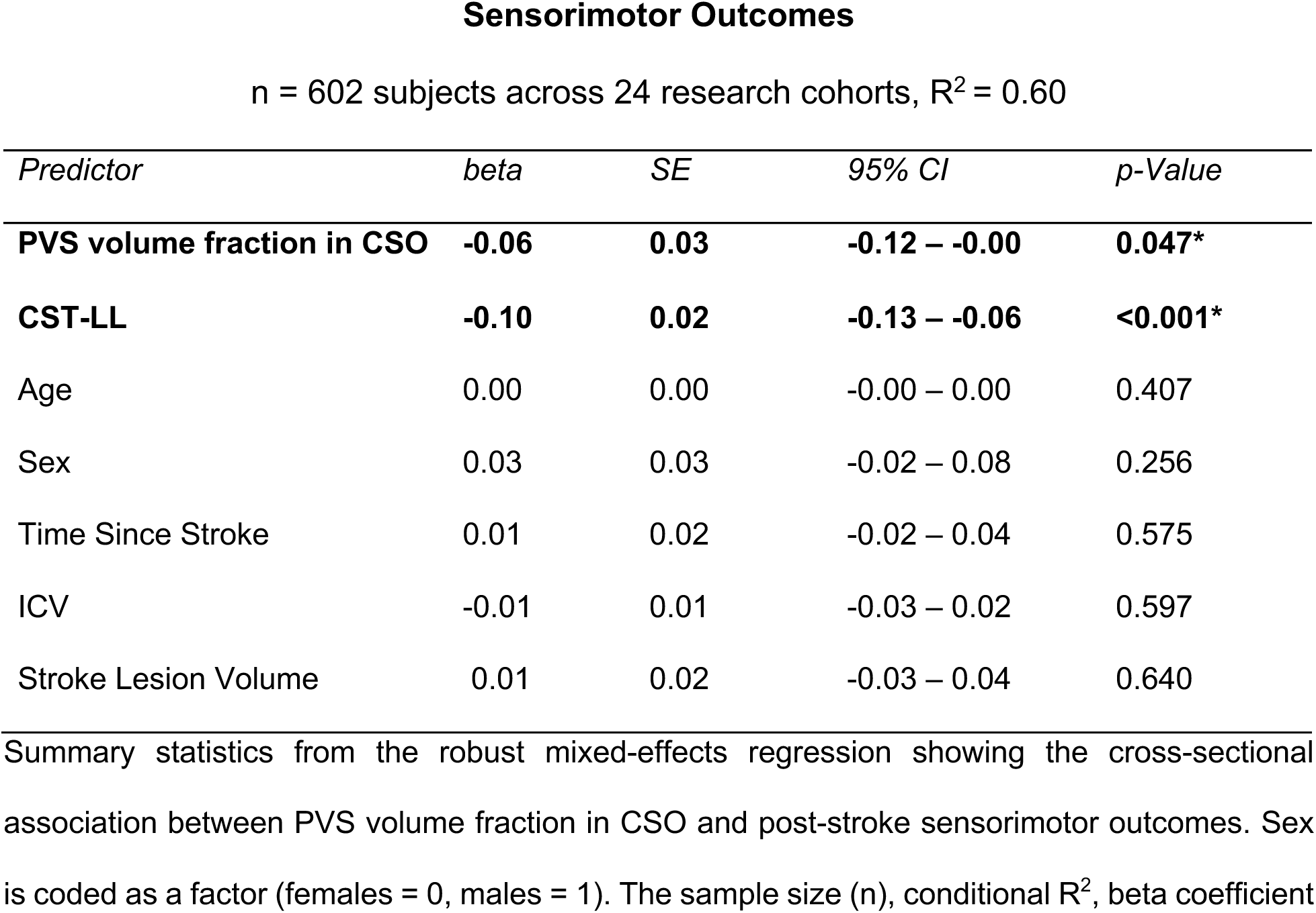
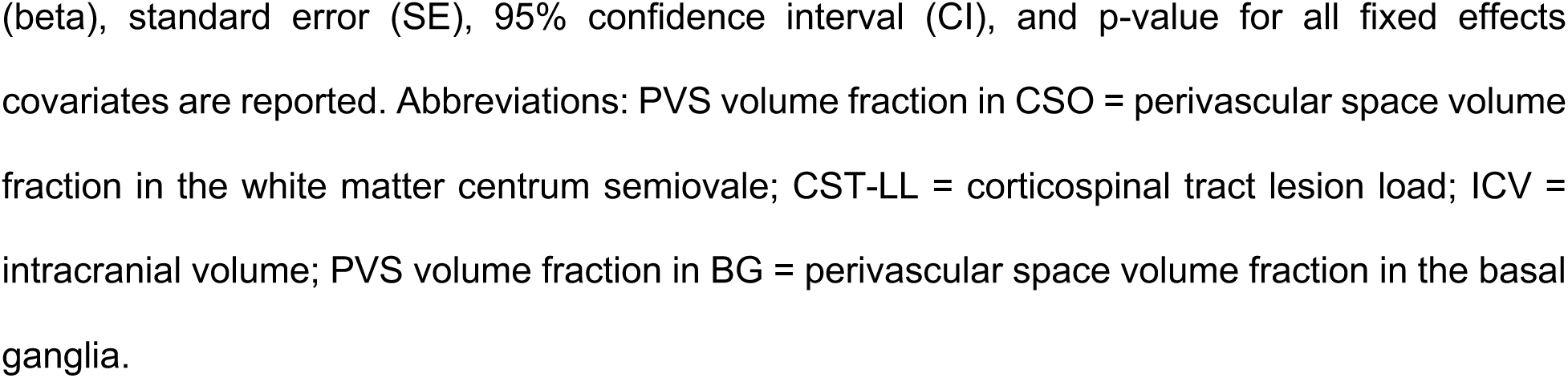
Association Between PVS Volume Fraction and Post-Stroke Sensorimotor Outcomes.

| *Predictor* | *beta* | *SE* | *95% CI* | *p-Value* |
| --- | --- | --- | --- | --- |
| **PVS volume fraction in CSO** | **-0.06** | **0.03** | **-0.12 – -0.00** | **0.047\*** |
| **CST-LL** | **-0.10** | **0.02** | **-0.13 – -0.06** | **<0.001\*** |
| Age | 0.00 | 0.00 | -0.00 – 0.00 | 0.407 |
| Sex | 0.03 | 0.03 | -0.02 – 0.08 | 0.256 |
| Time Since Stroke | 0.01 | 0.02 | -0.02 – 0.04 | 0.575 |
| ICV | -0.01 | 0.01 | -0.03 – 0.02 | 0.597 |
| Stroke Lesion Volume | 0.01 | 0.02 | -0.03 – 0.04 | 0.640 |

Summary statistics from the robust mixed-effects regression showing the cross-sectional association between PVS volume fraction in CSO and post-stroke sensorimotor outcomes. Sex is coded as a factor (females = 0, males = 1). The sample size ( $n$ ), conditional $R^2$ , beta coefficient
(beta), standard error (SE), 95% confidence interval (CI), and p-value for all fixed effects covariates are reported. Abbreviations: PVS volume fraction in CSO = perivascular space volume fraction in the white matter centrum semiovale; CST-LL = corticospinal tract lesion load; ICV = intracranial volume; PVS volume fraction in BG = perivascular space volume fraction in the basal ganglia.

### Higher Burden of Deep, Periventricular WMH, Brain-PAD, Older Age and Male Sex are Associated with Larger PVS Volume Fraction in the CSO

PVS volume fraction in the CSO was associated with a higher burden of deep WMH (b = 0.25, 95% CI = 0.19 - 0.31, p <0.001), periventricular WMH (b = 0.16, 95% CI = 0.09 - 0.23, p = <0.001) and brain-PAD (b = 0.09, 95% CI = 0.05 - 0.13, p <0.001). In addition, demographic factors such as older age (b = 0.01, 95% CI = 0.01 - 0.01, p <0.001) and male sex (b = 0.10, 95% CI = 0.03 - 0.17, p = 0.003) were also significantly associated with larger PVS volume fraction in the CSO (Table III). However, time since stroke, stroke lesion volume, CST-LL and ICV were not associated.

**Table III:**
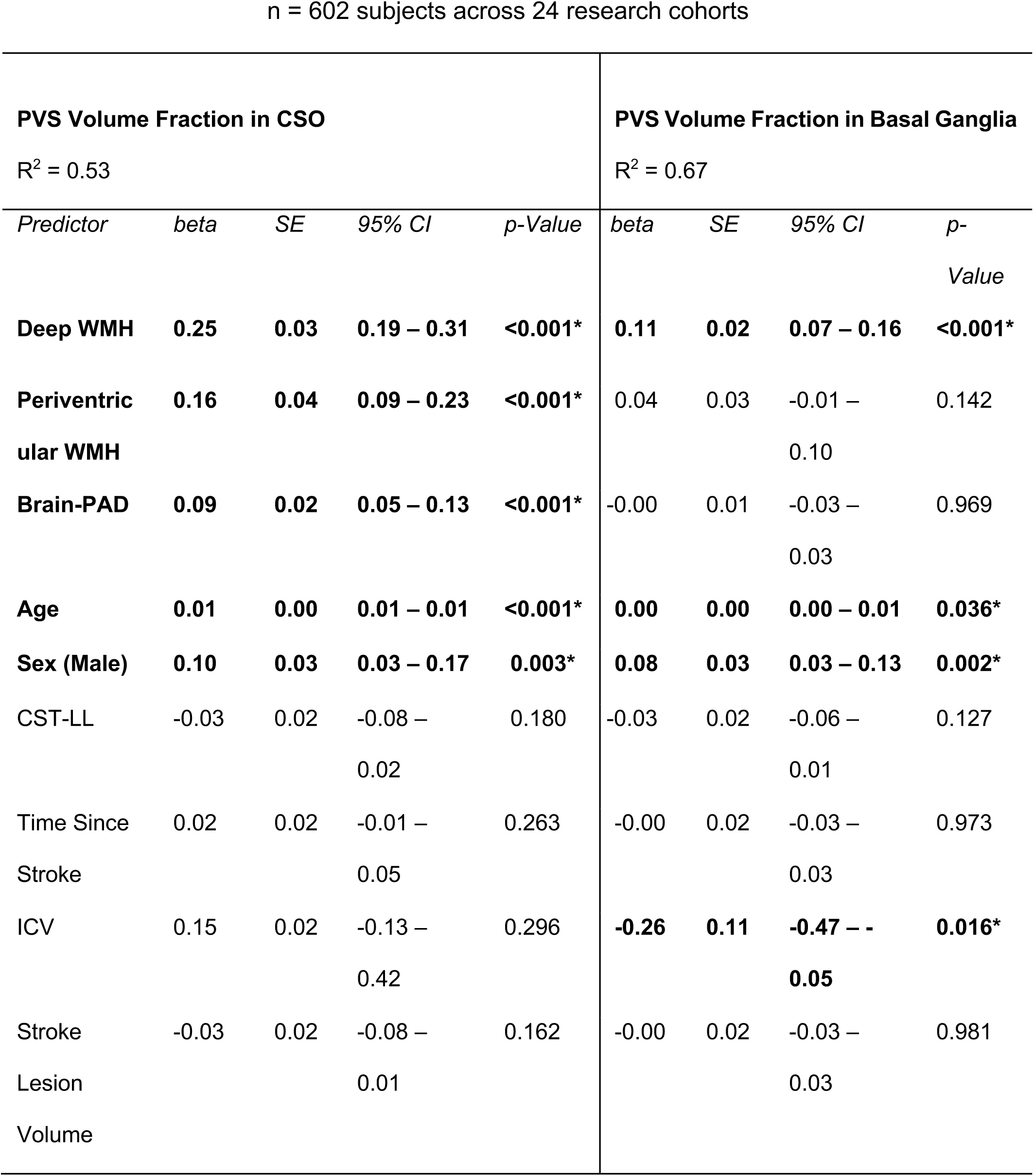
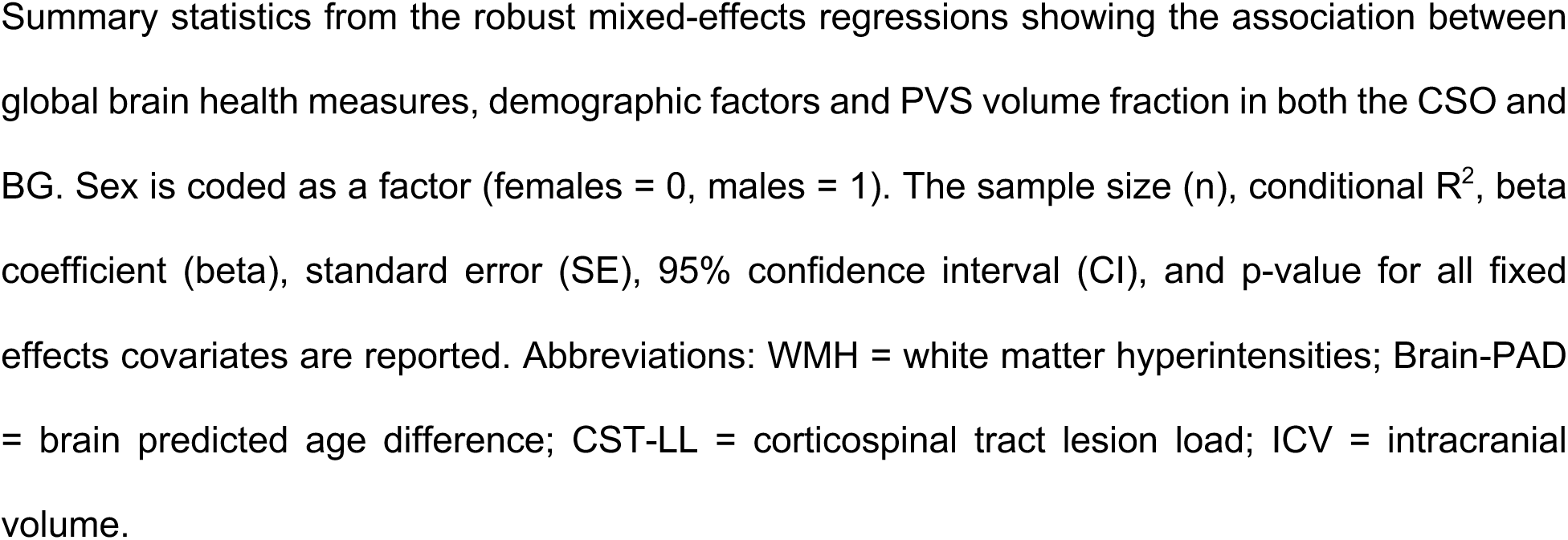
Association between Global Brain Health Measures and PVS Volume Fraction in the CSO and Basal Ganglia.

n = 602 subjects across 24 research cohorts
| **PVS Volume Fraction in CSO**<br>R<sup>2</sup> = 0.53 |  |  |  |  | **PVS Volume Fraction in Basal Ganglia**<br>R<sup>2</sup> = 0.67 |  |  |  |
| --- | --- | --- | --- | --- | --- | --- | --- | --- |
| *Predictor* | *beta* | *SE* | *95% CI* | *p-Value* | *beta* | *SE* | *95% CI* | *p-Value* |
| **Deep WMH** | **0.25** | **0.03** | **0.19 – 0.31** | **<0.001\*** | **0.11** | **0.02** | **0.07 – 0.16** | **<0.001\*** |
| **Periventricular WMH** | **0.16** | **0.04** | **0.09 – 0.23** | **<0.001\*** | 0.04 | 0.03 | -0.01 – 0.10 | 0.142 |
| **Brain-PAD** | **0.09** | **0.02** | **0.05 – 0.13** | **<0.001\*** | -0.00 | 0.01 | -0.03 – 0.03 | 0.969 |
| **Age** | **0.01** | **0.00** | **0.01 – 0.01** | **<0.001\*** | **0.00** | **0.00** | **0.00 – 0.01** | **0.036\*** |
| **Sex (Male)** | **0.10** | **0.03** | **0.03 – 0.17** | **0.003\*** | **0.08** | **0.03** | **0.03 – 0.13** | **0.002\*** |
| CST-LL | -0.03 | 0.02 | -0.08 – 0.02 | 0.180 | -0.03 | 0.02 | -0.06 – 0.01 | 0.127 |
| Time Since Stroke | 0.02 | 0.02 | -0.01 – 0.05 | 0.263 | -0.00 | 0.02 | -0.03 – 0.03 | 0.973 |
| ICV | 0.15 | 0.02 | -0.13 – 0.42 | 0.296 | **-0.26** | **0.11** | **-0.47 – -0.05** | **0.016\*** |
| Stroke Lesion Volume | -0.03 | 0.02 | -0.08 – 0.01 | 0.162 | -0.00 | 0.02 | -0.03 – 0.03 | 0.981 |

Summary statistics from the robust mixed-effects regressions showing the association between global brain health measures, demographic factors and PVS volume fraction in both the CSO and BG. Sex is coded as a factor (females = 0, males = 1). The sample size (n), conditional $R^2$ , beta coefficient (beta), standard error (SE), 95% confidence interval (CI), and p-value for all fixed effects covariates are reported. Abbreviations: WMH = white matter hyperintensities; Brain-PAD = brain predicted age difference; CST-LL = corticospinal tract lesion load; ICV = intracranial volume.

### Higher Burden of Deep WMH, Intracranial Volume, Older Age and Male Sex are Associated with Larger PVS Volume Fraction in the Basal Ganglia

We found that a higher burden of deep WMH (b = 0.11, 95% CI = 0.07 – 0.16, p <0.001), older age (b = 0.00, 95% CI = 0.00 – 0.01, p = 0.036), male sex (b = 0.08, 95% CI = 0.03 – 0.13, p = 0.002) and ICV (b = -0.26, 95% CI = -0.47 - -0.05, p = 0.016) were significantly associated with BG PVS volume fraction (Table III). However, periventricular WMH, brain-PAD, time since stroke, stroke lesion volume and CST-LL were not.

## DISCUSSION

### Larger PVS Volume Fraction in the CSO is Associated with Worse Post-Stroke Sensorimotor Outcomes

In this study, we investigated the cross-sectional association between PVS volume fraction and post-stroke sensorimotor outcomes in a large, multi-site, retrospective cohort of stroke subjects, after controlling for age, sex, time since stroke, CST-LL, ICV and stroke lesion volume. We found that a larger PVS volume fraction in the CSO is associated with worse post-stroke sensorimotor outcomes. To our knowledge, this is the first study to show a relationship between PVS volume fraction and post-stroke sensorimotor outcomes, above and beyond other lesion-based biomarkers of post-stroke motor outcomes like CST-LL or lesion volume.^2,46^ PVS enlargement is a marker of CSVD and may signal poor overall vascular brain health after stroke.^21^ An important finding of this study is that PVS does not appear to be associated with any lesion or stroke-specific factors, such as lesion volume, CST-LL, or time since stroke, suggesting that PVS may occur independently of lesion damage, and that PVS and lesion damage (specifically, CST-LL) may independently impact sensorimotor damage through different mechanistic pathways.

In this cohort, it is possible that PVS enlargement was a result of preexisting or concurrent CSVD, or increased following stroke.^11^ Several possible mechanisms of PVS enlargement exist. First, enlargement of PVS could be attributed to inflammation, which is a common physiological response after stroke.^14,47^ Inflammation may lead to increased oxygen consumption and reduced blood flow, and inflammatory markers may be released into surrounding cerebral blood vessels, with pro-inflammatory cells often accumulating within the PVS.^48,49^ Inflammatory cells can cause breakdown of the blood-brain barrier (BBB) and the extracellular matrix, thereby disturbing tightly connected endothelial cells, leading to increased leakage of fluid from blood vessels and subsequently, PVS enlargement.^14,50^ Second, cerebral hypoxia and hypoperfusion are recognized mechanisms following stroke.^47,51,52^ Both hypoxia and hypoperfusion also cause structural alterations in blood vessels and endothelial dysfunction.^19,53^ Endothelial dysfunction-related structural alterations may include enlargement of the vessel lumen and blood vessel stiffening, which are both precursors of PVS enlargement.^,54^

We found larger PVS volume fraction in the CSO to be associated with post-stroke sensorimotor impairment. As described previously, PVS enlargement is a marker of CSVD.^21^ Pre-existing CSVD may be indicative of increased exposure of brain tissue to ischemia; cerebral ischemia may decrease the overall neurological reserve capacity, disrupt motor network architecture and cortical reorganization after stroke, impeding plasticity processes.^54,55,56^ Specifically, PVS enlargement in the CSO may be caused by impaired fluid drainage into the underlying white matter;^20^ widespread white matter microstructural damage and disruption of projection fibers that communicate with descending motor pathways^3,57^ may lead to subsequent motor neuron injury and present as sensorimotor impairment.^58, 83^

On the other hand, we did not find a significant relationship between basal ganglia PVS volume fraction and post-stroke sensorimotor impairment. PVS enlargement in the basal ganglia has been associated with motor dysfunction in neurodegenerative diseases like Parkinson’s Disease.^59,60^ Glymphatic dysfunction and inflammatory pathophysiological processes involved in basal ganglia PVS enlargement may also concurrently damage dopaminergic neurons in the substantia nigra. It is therefore possible that basal ganglia PVS enlargement may be associated with severity and progression of extrapyramidal motor control symptoms (e.g., rigidity, resting tremor, gait abnormalities in Parkinson’s^61^) rather than direct motor impairment as a result of stroke. Additionally, underlying cerebral amyloid angiopathy (CAA) could be another potential explanation for the lack of association between BG PVS volume fraction and sensorimotor outcomes. CAA is a distinctive pathology of CSVD^12^ which is indicative of aging and neurodegeneration^12^ and characterized by the deposition of beta-amyloid in the walls of microvessels. CAA has been associated with a greater burden of CSO PVS^11,13^ and in this cohort, since larger CSO PVS but not BG PVS volume fraction was associated with sensorimotor outcomes, it is possible that individuals with stroke also had underlying CAA. While these may be possible explanations for the lack of association between BG PVS volume fraction and sensorimotor outcomes, future detailed investigation accounting for more stroke-specific measures such as the location and type of stroke (e.g. using TOAST classification) is required to confirm the independent involvement of CSO PVS enlargement in post-stroke motor impairment.

### Larger PVS Volume Fraction is Associated with Measures of Poorer Global Brain Health

Our secondary analysis revealed that a larger PVS volume fraction in both CSO and BG is associated with a higher burden of measures of whole-brain vascular integrity (i.e., WMHs). Early studies in the stroke population demonstrated a stronger association between PVS enlargement in the basal ganglia and deep or periventricular WMHs.^22,62,63^ More recently, in two separate large cohorts of acute ischemic stroke patients, the presence of more severe deep and periventricular WMHs was associated with greater PVS enlargement in the CSO and BG.^64,65,66^ Our findings are in line with this observation. PVS enlargement and the formation of both deep and periventricular WMHs may share similar pathophysiological mechanisms, such as demyelination. Release of inflammatory cells due to underlying CSVD may trigger demyelination and tissue loss surrounding the perivascular compartment.^11,67,68^ Demyelination may lead to axonal damage, characterized by the loosening and eventual loss of white matter fibers, which could lead to enlargement of PVS,^57,67^ as well as the formation of deep and periventricular WMHs.^69^ This may explain our findings, which show that a higher burden of WMHs is associated with larger PVS volume fraction.^69,70,71^

Additionally, we found that higher brain-PAD (a measure of whole-brain structural integrity), indicating older-appearing brains, is associated with larger PVS volume fraction in the CSO. Brain-PAD is calculated as the difference between an individual’s predicted brain age and their chronological age; positive brain-PAD values indicate older-appearing brains and negative values indicate younger-appearing brains.^6,43^ A recent study showed that in individuals with unilateral stroke, an older-appearing brain may be related to ventricular enlargement, smaller cortical thickness and surface area, and smaller subcortical volume, suggesting that brain-PAD represents measures of whole-brain structural atrophy after stroke.^6^ Similar measures of structural brain atrophy have been linked to PVS enlargement in stroke, but primarily in the basal ganglia. For example, researchers detected an association between brain atrophy (measured by enlargement of the superficial cortical gyri and deep ventricles) and basal ganglia PVS enlargement in a cohort of individuals with lacunar infarcts.^72^ Aging accelerates degeneration, deposition of β-amyloid plaques, and facilitates widespread neuronal dysfunction, which ultimately lead to reduced protein waste removal and a decline in the efficient exchange of nutrients between the CSF and brain parenchyma^73^ – all of which are precursors for PVS enlargement. This may be one potential explanation for the association between measures of structural atrophy such as brain-PAD and PVS enlargement in the CSO.^51,74,75^

Finally, we also report associations between larger PVS volume fraction in both the BG and CSO, and demographic factors such as older age and male sex. These findings support existing literature and suggest that visibility of enlarged PVS increases with age, and may also be sex-specific.^20,51,64^

Strengths of this study include the use of a large, diverse, multi-site retrospective dataset with neuroimaging and sensorimotor outcome data. In addition, the international working group providing standards for research into small vessel disease (STRIVE guidelines) highly recommend the use of quantitative PVS segmentation pipelines such as what we used here, as they may be more sensitive to detecting subtle associations.^76,77^ Some limitations of the study primarily stem from our use of a large retrospective dataset, which does not contain all the specific variables we would like to examine. For instance, we are unable to account for vascular risk factors (e.g. hypertension, type 2 diabetes mellitus, high cholesterol) due to this missing information across our dataset. Hypertension specifically has been strongly linked to PVS enlargement in stroke and may have a confounding effect on the relationship between PVS volume fraction and stroke sensorimotor outcomes.^77,78,79^ Future studies may account for vascular risk and other lifestyle factors when testing for associations between PVS volume fraction and post-stroke sensorimotor outcomes.^80^ Second, since this was a cross-sectional analysis, we only report a relationship between PVS volume fraction from a single time point and post-stroke sensorimotor *outcomes,* and not *sensorimotor recovery* over time. Future longitudinal studies may examine change in PVS volume fraction and sensorimotor outcomes to determine trajectories of post-stroke *sensorimotor recovery*; this may enhance our understanding of the extent to which vascular brain health may have a role in explaining inter-individual differences in sensorimotor recovery after stroke. Third, our dataset was primarily chronic and only a structural T1-weighted image was available to use for PVS segmentation. Although T1-weighted images are acceptable according to the STRIVE neuroimaging guidelines,^76,77^ an accompanying T2-weighted/ FLAIR image could improve the accuracy of the PVS segmentation pipeline.^14^ Future studies may utilize T2-weighted or FLAIR images for improved segmentation accuracy and incorporate a wider range of subjects across the stroke recovery continuum.

## CONCLUSION

Our findings demonstrate a novel cross-sectional association between larger PVS volume fraction in the CSO and worse post-stroke sensorimotor outcomes. This finding is important because it shows that in addition to CST-LL, PVS volume fraction may be another potential measure of vascular brain health that may account for inter-individual variability in post-stroke sensorimotor outcomes, and which may be a future target for personalized stroke rehabilitation interventions. Our finding that PVS impacts sensorimotor outcomes independent of lesion-based metrics suggests that PVS may reflect underlying age-and CSVD-related mechanisms, rather than stroke-specific events. Future longitudinal studies could demonstrate whether the severity of PVS enlargement at the time of stroke can predict worse sensorimotor outcomes after stroke, allowing PVS to be used as a prognostic neuroimaging-based biomarker of post-stroke sensorimotor prognosis.^77^

## Non-standard Abbreviations and Acronyms

CST-LL: Corticospinal Tract Lesion Load
GBH: Global Brain Health
CSVD: Cerebral Small Vessel Disease
PVS: Perivascular Space
BG: Basal Ganglia
CSO: White Matter Centrum Semiovale
WMH: White Matter Hyperintensities
Brain-PAD: Brain Predicted Age Difference
ENIGMA: Enhancing NeuroImaging Genetics through Meta-Analysis
ICV: Intracranial Volume

## SOURCES OF FUNDING

1. Stuti Chakraborty – Chan Division of Occupational Science and Occupational Therapy

2. Jeiran Choupan – NA

3. Octavio Marin-Pardo – NIH NINDS R01NS115845, Period: 04/2020-03/2025

4. Mahir H. Khan – NIH NINDS R01NS115845, Period: 04/2020-03/2025

5. Giuseppe Barisano – NA

6. Bethany P. Tavenner – NA

7. Miranda R. Donnelly – NA

8. Aisha Abdullah – NA

9. Justin W. Andrushko -Canadian Institutes of Health Research, Michael Smith Foundation for Health Research, BC

10. Nerisa Banaj – NA

11. Michael R. Borich - NIH P2CHD086844, NIH U24NS107234

12. Lara A. Boyd – Canadian Institutes of Health Research (PI Boyd: PTJ-148535, MOP- 130269, MOP-106651), Centre for Progress in Stroke Recovery Grant, Canada

13. Cathrin M. Buetefisch – NIH R01NS090677, R21HD067906-01A1

14. Adriana B. Conforto - NIH R01 NS076348; IIEP-2250-14

15. Steven C. Cramer, MD - NIH UH3-NS121565, NIH U01 NS120910, National Institute of Neurological Disorders and Stroke Award: R01NS115845, Role: Co-I, California Rehabilitation Institute Research Grant 20214600, NIH U01 NS086872, NIH R01 HD062744, Role: Co-I, R01 HD095457, Department of Veterans Affairs Research Grant 36C24E21P0147, Role: Co-I

16. Martin Domin, Dr.rer.med – NA

17. Adrienne Dula A., PhD – Funding was made available by the Texas Legislature to the Lone Star Stroke Clinical Trial Network. Its contents are solely the responsibility of the authors and do not necessarily represent the official views of the Government of the United States or the State of Texas.

18. Jennifer K. Ferris – NA

19. Brenton Hordacre – NA

20. Steven A. Kautz - NIH P20 GM109040, VA Rehabilitation R&D 1IK6RX003075, VA Rehabilitation R&D 1I01RX001935

21. Neda Jahanshad - NIH R01AG059874 – PI, NIH R01MH117601 – MPI

22. Martin Lotze – NA

23. Kyle Nishimura – NA

24. Fabrizio Piras – Italiana Ministry of Health, Ricerca Corrente 2024

25. Kate P. Revill – NIH NINDS R01NS090677

26. Nicolas Schweighofer – NIH NINDS: R01NS115845, Role: Co-I, Period: 04/2020-03/2025

27. Surjo R. Soekadar - European Research Council (ERC NGBMI, 759370), Federal Ministry of Research and Education (BMBF SSMART, 01DR21025A), Deutsche Forschungsgemeinschaft (DFG SO932/7-1)

28. Shraddha Srivastava – NA

29. Sophia I. Thomopoulos – The ENIGMA-Stroke working group gratefully acknowledges support from the NIH Big Data to Knowledge (BD2K) award (U54 EB020403 to Paul Thompson). For a complete list of ENIGMA-related grant support please see here: http://enigma.ini.usc.edu/about-2/funding/. SIT was supported by NIH grants R01MH131806, R01MH129742 to PMT.

30. Daniela Vecchio – NA

31. Lars T. Westlye – The European Research Council under the European Union’s Horizon 2020 research and Innovation program (ERC StG Grant No. 802998).

32. Carolee J. Winstein – NIH/NICHD HD065438

33. George F. Wittenberg – Department of Veterans Affairs

34. Kristin A. Wong – NA

35. Paul M. Thompson - The ENIGMA-Stroke working group gratefully acknowledges support from the NIH Big Data to Knowledge (BD2K) award (U54 EB020403 to Paul Thompson). For a complete list of ENIGMA-related grant support please see here: http://enigma.ini.usc.edu/about-2/funding/. NIH R01MH131806, R01MH129742

36. Sook-Lei Liew – NIH NINDS R01NS115845

## SUPPLEMENTAL MATERIAL

Supplemental Methods Results

Table S1

Figure S1

