## Supplemental Material for "Larger perivascular space volume fraction is associated with worse post-stroke sensorimotor outcomes: An ENIGMA analysis"

### METHODS

#### PVS Segmentation

PVS segmentation masks were obtained from the automated segmentation pipeline described in the methods, which were visually inspected for quality with 5% of each site randomly inspected for segmentation accuracy. The Frangi filter threshold was chosen to ensure accurate whole-brain PVS segmentation across all sites and was set at 0.001 for BG PVS and 0.0002 for CSO PVS, respectively. However, since this dataset did not have an accompanying T2-weighted or FLAIR image, it was not possible to correct for white matter hyperintensities (WMH) that may have been mistaken as PVS, leading to a possible overestimation of PVS. However, this overestimation would have been consistent across all individuals. In addition, WMH were separately estimated via the gold standard visual rating scale and accounted for in our secondary analysis models (Models 2 and 3).

#### Comparison of automated versus manual PVS segmentation:

Visual ratings based on manually counting the number of PVS by experts is a gold-standard for comparison of automated PVS segmentation methods.[15,76] Visual ratings were determined for PVS in the BG and CSO by an expert neurologist (G.B.), using a 4-point rating scale. Here, a score of 0 indicated no PVS; 1 indicated between 1–10 PVS; 2 indicated between 11–20 PVS; 3 indicated between 21–40 PVS; and 4 indicated >40 PVS.[21] Additionally, the highest number of visible PVS appearing on a slice was manually counted by an expert (G.B.) and recorded. We used robust mixed effects regressions to test the association between automatically segmented PVS volume fraction, manual PVS count and manual PVS visual rating scores from both the BG and CSO. Age and sex were included as fixed effects covariates, with site as a random effect in the regression models. PVS volume fraction in the BG was significantly associated with both manual PVS count in the BG (b = 0.41, 95% CI = 0.29 – 0.52, p < 0.001) and manual PVS visual rating score in the BG (b = -0.15, 95% CI = -0.27 - -0.03, p < 0.001). However, PVS volume fraction in the CSO was significantly associated only with manual PVS count in the CSO (b = 0.25, 95% CI = 0.07 – 0.43, p = 0.005) (Supplementary Table I).

**Statistical Analyses:**

**Model 1:**

$$
\begin{aligned}
Sensorimotor\ Outcome \sim\ & PVS\ Volume\ Fraction\ in\ CSO + PVS\ Volume\ Fraction\ in\ BG \\
& + Age + Sex + Time\ Since\ Stroke + Corticospinal\ Tract\ Lesion\ Load \\
& + Intracranial\ Volume + Stroke\ Lesion\ Volume + (1|Research\ Cohort)
\end{aligned}
$$

**Model 2:**

$$
\begin{aligned}
PVS\ Volume\ Fraction\ in\ CSO \sim\ & Deep\ WMH + Periventricular\ WMH + brainPAD \\
& + Age + Sex + Time\ Since\ Stroke + Corticospinal\ Tract\ Lesion\ Load \\
& + Intracranial\ Volume + Stroke\ Lesion\ Volume + (1|Research\ Cohort)
\end{aligned}
$$

**Model 3:**

$$PVS\ Volume\ Fraction\ in\ BG \sim Deep\ WMH\ +\ Periventricular\ WMH\ + brainPAD$$
$$+\ Age\ +\ Sex\ +\ Time\ Since\ Stroke\ +\ Corticospinal\ Tract\ Lesion\ Load$$
$$+\ Intracranial\ Volume\ +\ Stroke\ Lesion\ Volume +\ (1|Research\ Cohort)$$

### RESULTS

**Supplementary Table I: Relationship between PVS Volume Fraction, Manual PVS Count and Manual PVS Visual Rating Scores in the BG and CSO**

| **PVS Volume Fraction in BG** | | | | |
|---|---|---|---|---|
| n = 600 subjects across 24 research cohorts, $R^2$ = 0.70 | | | | |
| *Predictor* | *beta* | *SE* | *95% CI* | *p-Value* |
| Age | 0.00 | 0.00 | 0.00 – 0.01 | **0.001** |
| Sex | 0.03 | 0.02 | -0.01 – 0.07 | 0.141 |
| Manual PVS Count | 0.41 | 0.06 | 0.29 – 0.52 | **<0.001*** |
| Manual PVS Visual Rating Score | -0.15 | 0.06 | -0.27 – -0.03 | **0.017*** |
| **PVS Volume Fraction in CSO** | | | | |
| n = 600 subjects across 24 research cohorts, $R^2$ = 0.31 | | | | |
| *Predictor* | *beta* | *SE* | *95% CI* | *p-Value* |
| Age | 0.01 | 0.00 | 0.01 – 0.02 | **<0.001*** |
| Sex | 0.09 | 0.03 | 0.03 – 0.16 | **0.005*** |
| Manual PVS Count | 0.25 | 0.09 | 0.07 – 0.43 | **0.005*** |

| | | | | |
|---|---|---|---|---|
| Manual PVS Visual Rating Score | 0.22 | 0.14 | -0.05 – 0.50 | 0.114 |

Summary statistics from the robust mixed-effects regressions showing the association between PVS volume fraction, manual PVS count and manual PVS visual rating score in both the BG and CSO. Sex is coded as a factor (females = 0, males = 1). The sample size (n), conditional $R^2$, beta coefficient (beta), standard error (SE), 95% confidence interval (CI), and p-value for all fixed effects covariates are reported. Abbreviations: PVS volume fraction in BG = perivascular space volume fraction in the basal ganglia; PVS volume fraction in CSO = perivascular space volume fraction in the white matter centrum semiovale.

**Supplementary Table II: Summary of Research Cohort Characteristics and Sensorimotor Outcome Measures**

**Research Cohort Characteristics and Sensorimotor Outcome Measures**

n = 602 subjects across 24 research cohorts

| *Research Cohort* | *Country* | *n (F/M)* | *Sensorimotor Measure* |
|---|---|---|---|
| 1 | USA | 33 (10/23) | FMA-UE |
| 2 | USA | 11(5/6) | FMA-UE |
| 3 | USA | 12(5/7) | FMA-UE |
| 4 | Germany | 16 (5/11) | FMA-UE |
| 5 | USA | 23(10/13) | FMA-UE |

| | | | |
|---|---|---|---|
| 6 | Norway | 90(25/65) | NIHSS |
| 7 | USA | 14(5/9) | FMA-UE |
| 8 | Germany | 12(3/9) | Motricity Index |
| 9 | USA | 17(10/7) | MRC |
| 10 | USA | 24(6/18) | FMA-UE |
| 11 | USA | 25(7/18) | FMA-UE |
| 12 | USA | 31(8/23) | FMA-UE |
| 13 | USA | 14(6/8) | FMA-UE |
| 14 | Brazil | 14(6/8) | FMA-UE |
| 15 | Italy | 67(27/40) | Barthel Index |
| 16 | Italy | 35(19/16) | Barthel Index |
| 17 | Brazil | 29(15/14) | FMA-UE |
| 18 | USA | 3(0/3) | FMA-UE |
| 19 | USA | 7(1/6) | FMA-UE |
| 20 | Australia | 36(11/25) | FMA-UE |

| | | | |
|---|---|---|---|
| 21 | Canada | 33(13/20) | FMA-UE |
| 22 | USA | 19(10/9) | NIHSS |
| 23 | USA | 10(6/4) | NIHSS |
| 24 | USA | 27(11/16) | FMA-UE |

Summary of research cohort characteristics and sensorimotor outcome measures. For each research cohort, the country, total number of subjects by sex (female/ male) and primary sensorimotor outcome measure used to calculate the sensorimotor outcome scores are listed. Abbreviations: F = Female; M = Male; FMA-UE = Fugl-Meyer Assessment of the Upper Extremity; MRC = Medical Research Council Scale for Muscle Strength; NIHSS = National Institutes of Health Stroke Scale.

**Additional exploratory analysis:**

Since male sex was significantly associated with PVS volumes in both the BG and white matter CSO, we tested for sex differences in PVS volumes from each of these regions, using unpaired t-tests. Males had a larger median PVS volume in both the BG and CSO compared to females. However, this difference was statistically significant only in the white matter CSO ($t = -2.534$, $df = 444.77$ $p = 0.001$), but not in the BG ($t = -1.848$, $df = 473.5$, $p = 0.06$). We also found no differences in sensorimotor outcomes based on sex ($t = 0.529$, $df = 460.55$ $p = 0.597$). Supplementary Figure I (A and B) shows box plot comparisons of PVS volumes in the BG and CSO, by sex.

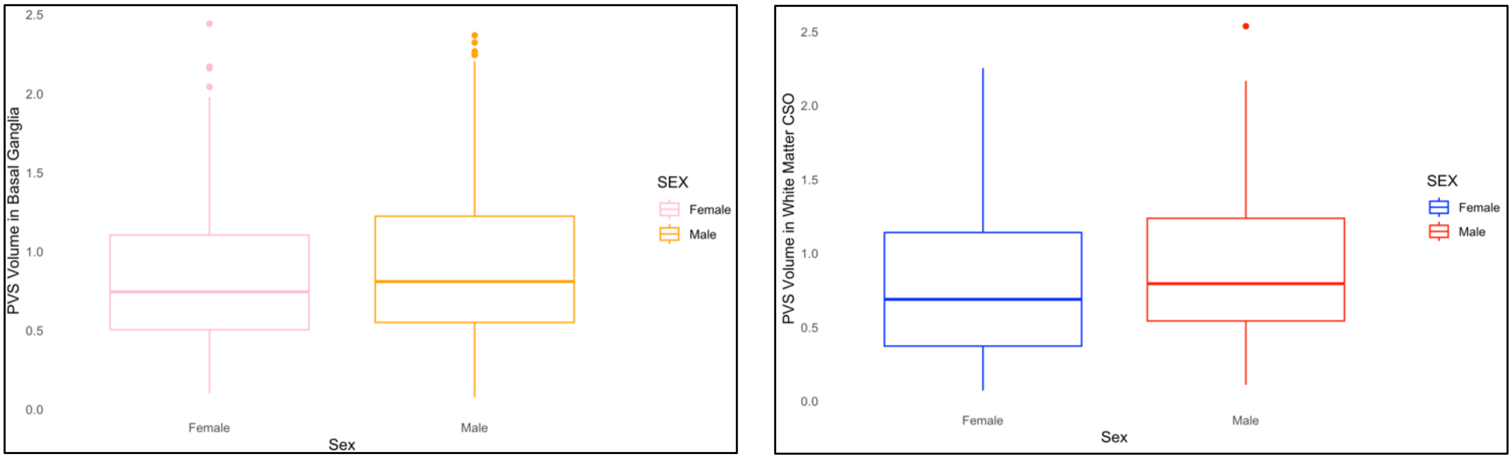


A B

Supplementary Figure I: Box plots comparing PVS volume fraction by sex in the BG (A) and CSO (B). Males demonstrated a higher median PVS volume fraction in both regions, but statistically significant differences are only seen in the CSO (t = -2.534, df = 444.77). p = 0.001
